# The Implementation Research Logic Model: A Method for Planning, Executing, Reporting, and Synthesizing Implementation Projects

**DOI:** 10.1101/2020.04.05.20054379

**Authors:** Justin D. Smith, Dennis H. Li, Miriam R. Rafferty

## Abstract

**Background:** Numerous models, frameworks, and theories exist for specific aspects of implementation research, including for determinants, strategies, and outcomes. However, implementation research projects often fail to provide a coherent rationale or justification for how these aspects are selected and tested in relation to one another. Despite this need to better specify the conceptual linkages between the core elements involved in projects, few tools or methods have been developed to aid in this task. The Implementation Research Logic Model (IRLM) was created for this purpose and to enhance the rigor and transparency of describing the often-complex processes of improving the adoption of evidence-based practices in healthcare delivery systems.

**Methods:** The IRLM structure and guiding principles were developed through a series of preliminary activities with multiple investigators representing diverse implementation research projects in terms of contexts, research designs, and implementation strategies being evaluated. The utility of the IRLM was evaluated in the course of a two-day training to over 130 implementation researchers and healthcare delivery system partners.

**Results:** Preliminary work with the IRLM produced a core structure and multiple variations for common implementation research designs and situations, as well as guiding principles and suggestions for use. Results of the survey indicated high utility of the IRLM for multiple purposes, such as improving rigor and reproducibility of projects; serving as a “roadmap” for how the project is to be carried out; clearly reporting and specifying how the project is to be conducted; and understanding the connections between determinants, strategies, mechanisms, and outcomes for their project.

**Conclusions:** The IRLM is a semi-structured, principles-guided tool designed to improve the specification, rigor, reproducibility, and testable causal pathways involved in implementation research projects. The IRLM can also aid implementation researchers and implementation partners in the planning and execution of practice change initiatives. Adaptation and refinement of the IRLM is ongoing, as is the development of resources for use and applications to diverse projects, to address the challenges of this complex scientific field.

## Background

In response to a call for addressing noted problems with transparency, rigor, openness, and reproducibility in biomedical research [1], the National Institutes of Health issued guidance in 2014 for improving the scientific rigor and reproducibility of the research it funds (https://www.nih.gov/research-training/rigor-reproducibility). The field of implementation science has similarly recognized a need for better specification with similar intent [2]. However, integrating the necessary conceptual elements of implementation research, which often involves multiple models, frameworks, and theories, is an ongoing challenge. A conceptually grounded organizational tool, specific to implementation research, could improve rigor and reproducibility while offering additional utility for the field.

This article describes the development and application of the Implementation Research Logic Model (IRLM). The model can be used with various types of implementation studies and at various stages of research, from planning to reporting and synthesizing completed studies. Examples of IRLMs are provided as Supplemental Materials for various common study designs and scenarios, including hybrid designs, studies involving multiple service delivery systems, and comparative implementation trials [3, 4]. Last, we describe preliminary use of the IRLM for training purposes and provide results from the post-training evaluation. An earlier version of this work was presented at the 2018 AcademyHealth/NIH Conference on the Science of Dissemination and Implementation in Health, and the abstract appeared in *Implementation Science* [5].

### Specification Challenges in Implementation Research

Having an imprecise understanding of what was done and why during implementation of a new innovation obfuscates identifying the factors responsible for successful implementation and prevents learning from what contributed to failed implementation. Thus, improving the specification of phenomena in implementation research is necessary to inform our understanding of how implementation strategies work, for whom, under what determinant conditions, and on what implementation and clinical outcomes. One challenge is that implementation science has numerous models and frameworks (hereafter, “frameworks”) to describe, organize, and aid in understanding the complexity of changing practice patterns and integrating evidence-based health interventions across systems [6]. These frameworks typically address implementation determinants, implementation process, or implementation evaluation [7]. Although many frameworks incorporate two or more of these broad purposes, researchers often find it necessary to use more than one to describe the various aspects of an implementation research study. The conceptual connections and relationships between multiple frameworks are often difficult to describe and to link to theory [8].

Similarly, reporting guidelines exist for some of these implementation research components, such as strategies [9] and outcomes [10], as well as for entire studies (i.e., Standards for Reporting Implementation Studies [11]); however, they generally help describe the individual components and not their interactions. To facilitate causal modeling [12], which can be used to elucidate mechanisms of change and the processes involved in both successful and unsuccessful implementation research projects, investigators must clearly define the relations among variables in ways that are testable with research studies [13]. Only then can we open the “black box” of how specific implementation strategies operate to predict outcomes.

### Logic Models

Logic models, a graphic depiction that presents the shared relationships among various elements of a program or study, have been used for decades in program development and evaluation [14] and are often required by funding agencies when proposing studies involving implementation [15]. Used to develop agreement among diverse stakeholders of the “what” and the “how” of proposed and ongoing projects, logic models have been shown to improve planning by highlighting theoretical and practical gaps, support the development of meaningful process indicators for tracking, and aid in both reproducing successful studies and identifying failures of unsuccessful studies [16]. They are also useful at other stages of research and for program implementation, such as organizing a project/grant application/study protocol, presenting findings from a completed project, and synthesizing the findings of multiple projects [17].

Logic models can also be used in the context of program theory to model explicitly how a clinical or preventive program (or an implementation strategy) contributes to a chain of intermediate results and subsequently to the intended outcomes. Program theory specifies a Theory of Change—the central processes or drivers by which change comes about following a formal theory or tacit understanding— and a Theory of Action, which explains how programs/strategies are constructed to activate the Theory of Change. Inherent within program theory is causal chain modeling. In implementation research, Fernandez et al. [18] applied mapping methods to implementation strategies to postulate the ways in which changes to the system affect downstream implementation and clinical outcomes. Their work presents an implementation mapping logic model based on Proctor et al. [19, 20], which is focused primarily on the selection of implementation strategy(s) rather than a complete depiction of the conceptual model linking all implementation research elements (i.e., determinants, strategies, mechanisms of action, implementation outcomes, clinical outcomes) in the detailed manner we describe in this article.

### Development of the IRLM

The development of the IRLM occurred through a series of case applications. This began with a collaboration between investigators at the Center for Prevention Implementation Methodology at Northwestern University and the Shirley Ryan AbilityLab in which the IRLM was used in the study of implementing a new model of patient care in a new physical space [21]. Next, the IRLM was used in the first 6 months of three already-funded implementation research projects to plan for and describe the prospective implementation research aspects of the trials, and an ongoing randomized roll-out implementation trial of the Collaborative Care Model for depression management [22]. It was also applied in the later stages of a nearly completed implementation research project testing two implementation strategies for implementing a family-based obesity management intervention in pediatric primary care to describe what had occurred over the course of the three-year trial [23]. Last, in a two-day training hosted by the Implementation Science Coordination, Consultation, and Collaboration Initiative at Northwestern University in October 2019, the IRLM was used as a training tool with all 65 grantees of NIH-funded planning project grants funded as part of the Ending the HIV Epidemic initiative [24]. Results of a survey from the ISC^3^I participants are reported in a later section of this article. From these preliminary activities, we identified a number of ways that the IRLM could be used, described in “Using the IRLM.”

## Methods

### The Implementation Research Logic Model

#### Structure

In developing the IRLM, we began with the common “pipeline” logic model format used by AHRQ, CDC, NIH, PCORI, and others [16]. This structure was chosen due to its familiarity to funders, investigators, readers, and reviewers. Although a number of characteristics of the pipeline logic model can be applied to implementation research studies, there is an overall misfit due to implementation research’s focusing on the systems that support adoption and delivery of health practices; involving multiple levels within one or more systems; and having its own unique terminology and frameworks [3, 25, 26]. We adapted the typical evaluation logic model to integrate existing implementation science frameworks as its core elements while keeping to the same aim of facilitating causal modeling.

The IRLM standard form is depicted in Figure 1 (Fillable PDF—Additional File A1). Variant formats are provided as Additional Files A2 to A5 for use with situations and study designs commonly encountered in implementation research, including comparative implementation studies (A2), studies involving multiple service contexts (A3), and implementation optimization designs (A4). A fourth variant includes a location to describe the EBP (A5), as preliminary work with the model indicated that some users desired a way to connect the intervention with the elements of the implementation research and disentangle it from the strategies.

**Figure 1.**
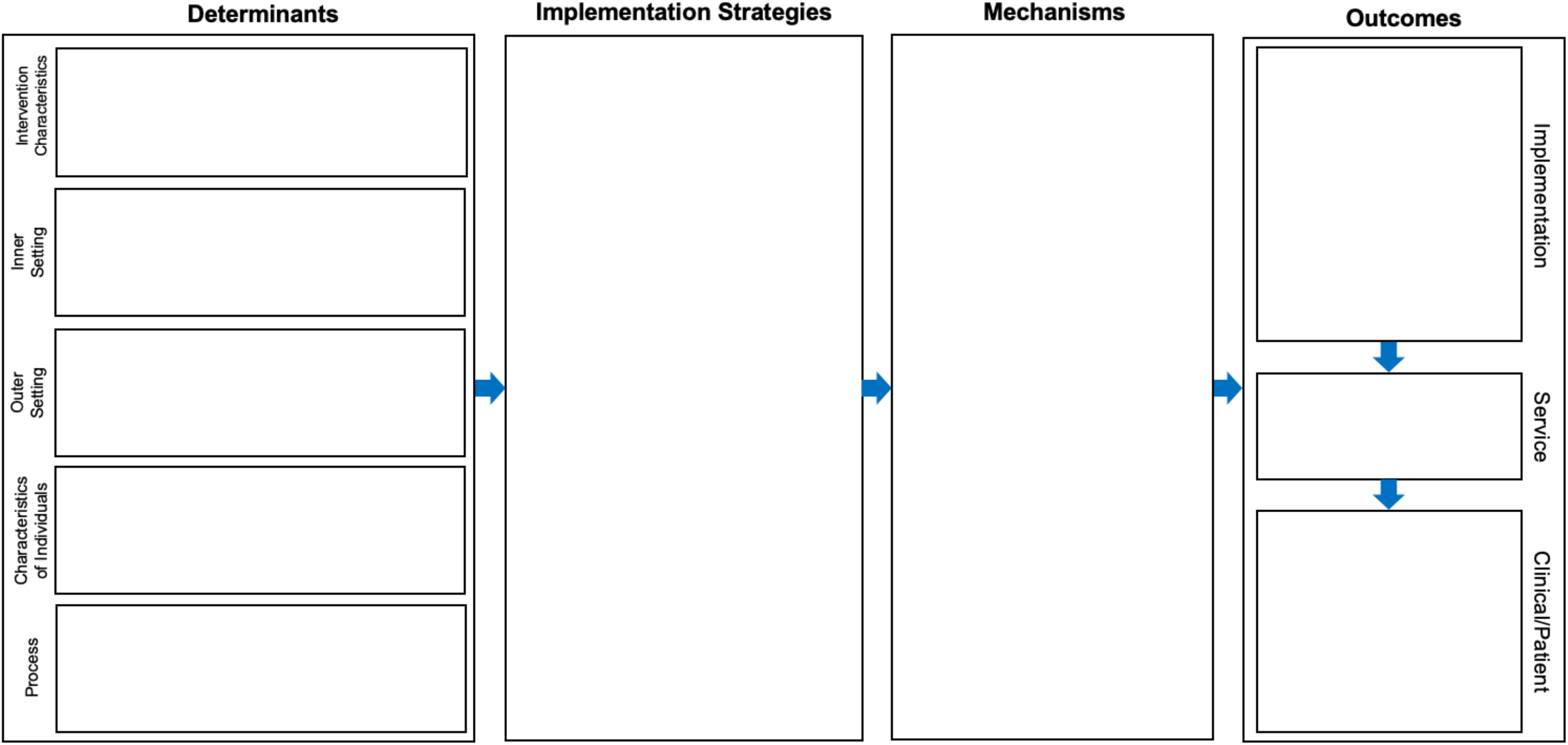
Implementation Research Logic Model (IRLM) Standard Form. *Notes*. Domain names in the determinants section were drawn from the Consolidated Framework for Implementation Research. The format of the outcomes section is from Proctor et al. 2011.

#### Core Elements and Theory

The IRLM specifies the relationships between determinants of implementation, implementation strategies, the mechanisms of action resulting from the strategies, and the implementation and clinical outcomes affected. These core elements are germane to every implementation research project in some way. Accordingly, the generalized theory of the IRLM posits that: (1) implementation strategies selected for a given evidence-based practice (EBP) are related to the implementation determinants (context-specific barriers and facilitators); (2) strategies work through specific mechanisms of action to change the context or the behaviors of those within the context; and (3) implementation outcomes are the proximal impacts of the strategy and its mechanisms, which then relate to the clinical outcomes of the EBP. Articulated in part by others [9, 12, 20, 27, 28], this causal pathway theory is largely explanatory and details the Theory of Change and the Theory of Action of the implementation strategies in a single model. The EBP Theory of Action can also be displayed within a modified IRLM (see Additional File A4). We now briefly describe the core elements and discuss conceptual challenges in how they relate to one another and to the overall goals of implementation research.

#### Determinants

Determinants of implementation are factors that might prevent or enable implementation (i.e., barriers and facilitators). Determinants may act as moderators, “effect modifiers,” or mediators, thus indicating that they are links in a chain of causal mechanisms [12]. Common determinant frameworks are the Consolidated Framework for Implementation Research (CFIR) [29] and the Theoretical Domains Framework [30].

#### Implementation strategies

Implementation strategies are supports, changes to, and interventions on the system to increase adoption of EBPs into usual care [31]. Consideration of determinants are commonly used when selecting and tailoring implementation strategies [27, 28, 32]. Providing the theoretical or conceptual reasoning for strategy selection is recommended [9]. The IRLM can be used to specify the proposed relationships between strategies and the other elements (determinants, mechanisms, and outcomes) and assists with considering, planning, and reporting all strategies in place during an implementation research project that could contribute to the outcomes and resulting changes

Because implementation research occurs within dynamic delivery systems with multiple factors that determine success or failure, the field has experienced challenges identifying consistent links between individual barriers and specific strategies to overcome them. For example, Waltz, Powell, Fernandez, Abadie, and Damschroder [28] attempted to use the Expert Recommendations for Implementing Change (ERIC) compilation of strategies [31] to determine which would best address contextual barriers identified by CFIR. An online CFIR–ERIC matching process completed by implementation researchers and practitioners resulted in a large degree of heterogeneity and few consistent relationships between barrier and strategy, meaning the relationship is rarely one-to-one (e.g., a single strategy is often is linked to multiple barriers; more than one strategy might be needed to address a single barrier). Moreover, when implementation outcomes are considered, researchers often find that to improve one outcome, more than one contextual barrier likely needs to be addressed, which might in turn require one or more strategies.

Frequently the reporting of implementation research studies focuses on the strategy or strategies that were introduced for the research study, without due attention to other strategies already used in the system or additional strategies that might be needed to implement the target strategy. The IRLM allows for comprehensive specification of all introduced and present strategies, as well as their changes (adaptations, additions, discontinuations) during the project.

#### Mechanisms of action

Mechanisms of action are processes or events through which an implementation strategy operates to affect desired implementation outcomes [12]. The mechanism can be a change in a determinant, a proximal implementation outcome, an aspect of the implementation strategy itself, or a combination of these in a multiple-intervening-effect model. An example of a causal process might be using training and fidelity monitoring strategies to improve delivery agents’ knowledge and self-efficacy about the EBP in response to knowledge-related barriers in the service delivery system. This could result in raising their acceptability of the EBP, increase the likelihood of adoption, improve the fidelity of delivery, and lead to sustainment.

Relatively few implementation studies formally test mechanisms of action, but this area of investigation has received significant attention more recently as the necessity to understand how strategies operate grows in the field [32-34].

#### Outcomes

Implementation outcomes are the effects of deliberate and purposive actions to implement new treatments, practices, and services [20]. They can be indicators of implementation processes, or key intermediate outcomes in relation to service, or target clinical outcomes. Glasgow et al. [35-37] describe the interrelated nature of implementation outcomes as occurring in a logical, but not necessarily linear, sequence of adoption by a delivery agent, delivery of the innovation with fidelity, reach of the innovation to the intended population, and sustainment of the innovation over time. The combined impact of these nested outcomes, coupled with the size of the effect of the EBP, determines the population or public health impact of implementation [35]. Outcomes earlier in the sequence can be conceptualized as mediators and mechanisms of strategies on later implementation outcomes. Specifying which strategies are theoretically intended to affect which outcomes, through which mechanisms of action, is crucial for improving the rigor and reproducibility of implementation research and to testing theory.

## Using the Implementation Research Logic Model

### Guiding Principles

One of the critical insights from our preliminary work was that use of the IRLM should be guided by a set of principles rather than governed by rules. These principles are intended to be flexible both to allow for adaptation to the various types of implementation studies and evolution of the IRLM over time and to address concerns in the field of implementation science regarding specification, rigor, reproducibility, and transparency of design and process [5].

### Principle 1: Strive for Comprehensiveness

Comprehensiveness increases transparency; can improve rigor; and allows for better understanding of alternative explanations to the conclusions drawn, particularly in the presence of null findings for an experimental design. Thus, all relevant determinants, implementation strategies, and outcomes should be included in the IRLM.

### Determinants

Concerning determinants, the valence should be noted as being either a barrier, a facilitator, neutral, or variable by study unit. This can be achieved by simply adding plus (+) or minus (–) signs for facilitators and barriers, respectively, or by using a coding systems such as that developed by Damschroder et al. [38], which indicates the relative strength of the determinant on a scale: –2 (*strong negative impact*), –1 (*weak negative impact*), 0 (*neutral or mixed influence*), 1 (*weak positive impact*), 2 (*strong positive impact*). Use of such a coding system could yield better specification compared to using study-specific adjectives or changing the name of the determinant (e.g., greater relative priority, addresses patient needs, good climate for implementation). It is critical to include all relevant determinants and not simply limit reporting to those that are hypothesized to be related to the strategies and outcomes, as there are complex interrelationships between determinants.

### Implementation strategies

Implementation strategies should be reported in their entirety. When using the IRLM for planning a study, it is important to list all strategies in the system, including those already in use and those to be initiated for the purposes of the study, often in the experimental condition of the design. Second, strategies should be labeled to indicate whether they were (a) in place in the system prior to the study, (b) initiated prospectively for the purposes of the study (particularly for experimental study designs), (c) removed as a result of being ineffective or onerous, or (d) introduced during the study to address an emergent barrier or supplement other strategies because of low initial impact. This is relevant when using the IRLM for planning, as an ongoing tracking system, for retrospective application to a completed study, and in the final reporting of a study. There have been a number of processes proposed for tracking the use of and adaptations to implementation strategies over time [39, 40]. Each of these are more detailed than would be necessary for the IRLM, but the processes described provide a method for accurately tracking the temporal aspects of strategy use that fulfil the Comprehensiveness principle.

### Outcomes

Although most studies will indicate a primary implementation outcome, other outcomes are almost assuredly to be measured. Thus, they ought to be included in the IRLM. This guidance is given in large part due to the interdependence of implementation outcomes, such that adoption relates to delivery with fidelity, reach of the intervention, and potential for sustainment [35]. Similarly, the overall public health impact (defined as reach multiplied by the effect size of the intervention [37]) are inextricably tied to adoption, fidelity, acceptability, cost, etc. Although the study’s focus might be justifiably on only one or two implementation outcomes, the others are nonetheless relevant and should be specified and reported.

### Principle 2: Indicate Key Conceptual Relationships

Although the IRLM has a generalized theory (described earlier), there is a need to indicate the relationships between elements in a manner aligning with the specific theory of change for the study. Researchers ought to provide some form or notation to indicate these conceptual relationships using color-coding, superscripts, arrows, or a combination of the three. Such notations in the IRLM facilitate reference in text to the study hypotheses, tests of effects, causal chain modeling, and other forms of elaboration (see “Supporting Text and Resources”). We prefer the use of superscripts to color or arrows in grant proposals and articles for practical purposes, as colors can be difficult to distinguish, and arrows can obscure text and contribute to visual convolution. When presenting the IRLM using presentation programs (e.g., PowerPoint, Keynote), colors and arrows can be helpful, and animations can make these connections dynamic and sequential without adding to visual complexity. This principle could also prove useful in synthesizing across similar studies to build the science of tailored implementation, where strategies are selected based on the presence of specific combinations of determinants. As previously indicated [28], there is much work to be done in this area given.

### Principle 3: Specify Critical Study Design Elements

This critical element will vary by the study design (e.g., Hybrid effectiveness-implementation trial; observational; what subsystems are assigned to the strategies). This principle includes not only researchers but service systems and communities, whose consent is necessary to carry out any implementation design [3, 41, 42].

#### Primary outcome(s)

Indicate the primary outcome(s) at each level of the study design (i.e., clinician, clinic, organization, county, state, nation). The levels should align with the specific aims of a grant application or the stated objective of a research report. In the case of a process evaluation or an observational study including the RE-AIM evaluation components [37] or the Proctor et al. [20] taxonomy of implementation outcomes, the primary outcome may be the product of the conceptual or theoretical model used when *a priori* outcomes are not clearly indicated. We also suggest including downstream health services and clinical outcomes even if they are not measured, as these are important for understanding the logic of the study and the ultimate health-related targets.

#### For quasi/experimental designs

When quasi/experimental designs [3, 4] are used, the independent variable(s) (i.e., the strategies that are introduced or manipulated or that otherwise differentiate study conditions) should be clearly labeled. This is important for internal validity and for differentiating conditions in multi-arm studies.

#### For comparative implementation trials

In the context of comparative implementation trials [3, 4], a study of two or more competing implementation strategies are introduced for the purposes of the study (i.e., the comparison is not implementation- as-usual), and there is a need to indicate the determinants, strategies, mechanisms, and potentially outcomes that differentiate the arms (see Additional File A2). As comparative implementation can involve multiple service delivery systems, the determinants, mechanisms, and outcomes might also differ, though there must be at least one comparable implementation outcome. In our preliminary work applying the IRLM to a large-scale comparative implementation trial, we found that we needed to use an IRLM for each arm of the trial as it was not possible to use a single IRLM because the strategies being tested occurred across two delivery systems and strategies were very different, by design. This is an example of the flexible use of the IRLM.

#### For implementation optimization designs

A number of designs are now available that aim to test processes of optimizing implementation. These include factorial, Sequential Multiple Assignment Randomized Trial (SMART) [43], adaptive [44], and roll- out implementation optimization designs [45]. These designs allow for: a) building time-varying adaptive implementation strategies based on the order in which components are presented [43]; b) evaluating the additive and combined effects of multiple strategies [43, 46]; and c) can incorporate data-driven iterative changes to improve implementation in successive units [44, 45]. The IRLM in Additional File A4 can be used for such designs.

#### Additional specification options

Users of the IRLM are allowed to specify any number of additional elements that may be important to their study. For example, one could notate those elements of the IRLM that have been or will be measured versus those that were based on the researcher’s prior studies or inferred from findings reported in the literature. Users can also indicate when implementation strategies differ by level or unit within the study. In large multisite studies, strategies might not be uniform across all units, particularly those strategies that already exist within the system. Similarly, there might be a need to increase the dose of certain strategies to address the relative strengths of different determinants within units.

### Using the IRLM for Different Purposes and Stages of Research

Commensurate with logic models more generally, the IRLM can be used for planning and organizing a project; carrying out a project (as a roadmap); reporting and presenting the findings of a completed project; and synthesizing the findings of multiple projects or of a specific area of implementation research, such as what is known about how learning collaboratives are effective within clinical care settings.

### Planning

When the IRLM is used for planning, the process of populating each of the elements often begins with the known parameter(s) of the study. For example, if the problem is improving the adoption and reach of a specific EBP within a particular clinical setting, the implementation outcomes and context, as well as the EBP, are clearly known. The downstream clinical outcomes of the EBP are likely also known. Working from the two “bookends” of the IRLM, the researchers and community partners and/or organization stakeholders can begin to fill in the implementation strategies that are likely to be feasible and effective and then posit conceptually derived mechanisms of action. In another example, only the EBP and primary clinical outcomes were known. The IRLM was useful in considering different scenarios for what strategies might be needed and appropriate to test the implementation of the EBP in different service delivery contexts. The IRLM was a tool for the researchers and stakeholders to work through these multiple options.

### Executing

When we used the IRLM to plan for the execution of funded implementation studies, the majority of the parameters were already proposed in the grant application. However, through completing the IRLM prior to the start of the study, we found that a number of important contextual factors had not been considered, additional implementation strategies were needed to complement the primary ones proposed in the grant, and mechanisms needed to be added and measured. At the time of award, mechanisms were not an expected component of implementation research projects as they will likely become in the future.

### Reporting

For another project, the IRLM was applied retrospectively to report on the findings and overall logic of the study. Because nearly all elements of the IRLM were known, we approached completion of the model as a means of showing what happened during the study and to accurately report the hypothesized relationships that we observed. These relationships could be formally tested using causal pathway modeling [12] or other path analysis approaches with one or more intervening variables [47].

### Synthesizing

In our preliminary work with the IRLM, we used it in each of the first three ways; the fourth (synthesizing) is ongoing within the National Cancer Institute’s Improving the Management of symPtoms during And Following Cancer

#### Treatment (IMPACT) research consortium

The purpose is to draw conclusions for the implementation of an EBP in a particular context (or across contexts) that are shared and generalizable to provide a guide for future research and implementation.

### Use of Supporting Text and Documents

While the IRLM provides a good deal of information about a project in a single visual, researchers often need to convey additional details about an implementation research study. We recommend making use of supporting text, tables, and figures to elaborate upon the IRLM in grant applications, reports, and articles. Some elements that will need elaboration are (a) preliminary data on the assessment and valence of implementation determinants; (b) implementation strategies being used or observed, using established reporting guidelines [9] and labeling conventions [31] from the literature; (c) hypothesized or tested causal pathways [12] ; (d) process, service, and clinical outcome measures, including the psychometric properties, method and timing of administration, respondents, etc.; (e) study procedures, including subject selection, assignment to (or observation of natural) study conditions, and assessment throughout the conduct of the study [4]; and (f) the implementation plan or process for following established implementation frameworks [48-50]. By utilizing superscripts, subscripts, and other notations within the IRLM, as previously suggested, it is easy to refer to (a) hypothesized causal paths in theoretical overviews and analytic plan sections; (b) planned measures for determinants and outcomes; and (c) specific implementation strategies in text, tables, and figures.

## Results

### Evidence of IRLM Utility and Acceptability

#### Testimonials

When we began development of the IRLM in collaboration with other research teams, we did not systematically collect outcomes, but we did ask for feedback on the usefulness of the IRLM and what it provided to their project. In the words of the Principal Investigator of one of the groups who used the IRLM for a recently funded study:

> “Completing the IRLM for our project helped both study arms systematically think through the steps needed to ultimately achieve the implementation outcomes. From the researcher standpoint, delineating the mechanistic pathways between each strategy and the outcomes creates a set of hypotheses that can be tested in the current study or in future research. For staff, identifying relevant determinants and selecting appropriate strategies to address those determinants helped inform the development of trainings and resources needed to successfully implement our intervention. This process, in turn, will inform how we design and scale out technical assistance in the future. For our staff with limited backgrounds in implementation research, there was a learning curve to understanding and completing the IRLM. We found breaking the project down into each element was helpful for their understanding. Once trained, they agreed the IRLM exercise was very useful for ensuring the comprehensiveness of the research plan.”

Similar opinions have been expressed by other research teams with whom we have worked.

#### Survey on the Utility of the IRLM

The IRLM was used as the foundation for a training in implementation research methods to a group of 65 planning projects awarded under the national Ending the HIV Epidemic initiative. One investigator (project director or co-investigator) and one implementation partner (i.e., partner in a community service delivery system) from each project were invited to attend a two-day in-person summit in Chicago, IL, in October 2019. One hundred and thirty-two participants attended, representing 63 of the 65 projects. A survey, which included demographics and questions pertaining to the Ending the HIV Epidemic, was sent to potential attendees prior to the summit, to which 129 individuals—including all 65 project directors, 13 co-investigators, and 51 implementation partners (62% Female)—responded. Those who indicated an investigator role (n=78) received additional questions about prior implementation research training (e.g., formal coursework, workshop, self-taught) and related experiences (e.g., involvement in a funded implementation project, program implementation, program evaluation, quality improvement) and the stage of their project (i.e., exploration, preparation, implementation, sustainment [49]). Approximately 6 weeks after the summit, 89 attendees (69%) completed a post-summit survey. Data from 42 investigators (65%) and 24 implementation partners who indicated having attended the training (N=66, 68.2% Female) were included in the following analyses.

Among the post-summit survey questions were 10 items related to the IRLM (see next paragraph), and one more generally about the logic of implementation research, each rated on a 4-point scale (0=*not at all*, 1=*a little*, 2=*moderately*, 3=*very much*).

Results were promising for the utility of the IRLM on the majority of the dimensions assessed. Respondents indicated that the IRLM was either moderately or very helpful in (a) improving the rigor and reproducibility (77.7%, M=3.05, *SD*=.885); (b) serving as a “roadmap” for how the project is to be carried out over time (74%, M=3.08, *SD*=.950); (c) clearly reporting and specifying how the project is to be conducted (67.8%, M=2.94, *SD*=.909); (d) understanding the connections between determinants, strategies, mechanisms, and outcomes (66.3%, M=2.92, *SD*=.957); (e) identifying gaps in the implementation research logic of their project (64.2%, M=2.86, *SD*=1.021); (f) deepening their knowledge of implementation science methods (62.9%, M=2.83, *SD*=.959); (g) planning the project (61.3%, M=2.82, *SD*=1.088); developing consensus and understanding of the project among diverse stakeholders involved (58.8%, M=2.75, *SD*=1.090); and (h) identifying gaps in new research questions or analyses (51.3%, M=2.54, *S*=1.032). They also indicated that the worksheets provided during the summit were helpful in completing the IRLM (74.1%, M=3.02, *SD*=.886). Overall, 77.6% (M=3.18, *SD*=.827) of respondents indicated that their knowledge on the logic of implementation research had increased either moderately or very much after the two-day training. At the time of the survey, when respondents were about 2.5 months into their one-year planning projects, 44.6% indicated that they had already been able to complete a full draft of the IRLM.

Additional analyses using one-way analysis of variance indicated no statistically significant differences in responses to the IRLM questions between investigators and implementation partners. However, three items approached significance: planning the project (*F*=2.460, *p*=.055); clearly reporting and specifying how the project is to be conducted (*F*=2.327, *p*=.066); and knowledge on the logic of implementation research (*F*=2.107, *p*=.091). In each case, scores were higher for the investigators compared to the implementation partners, suggesting that perhaps the knowledge gap in implementation research lay more in the academic realm than among community partners, who may not have a focus on research but whose day-to-day roles include the implementation of EBPs in the real world. Lastly, analyses using ordinal logistic regression did not yield any significant relationship between responses to the IRLM survey items and prior training (n=42 investigators who attended the training and completed the post-training survey), prior related research experience (n=42), and project stage of implementation (n=66). This suggests that the IRLM is a useful tool for both investigators and implementers with varying levels of prior exposure to implementation research concepts and across all stages of implementation research. As a result of this training, the IRLM is now a required element in the FY2020 Ending the HIV Epidemic Centers for AIDS Research/AIDS Research Centers Supplement Announcement released March 2020 [15].

### Resources for Using the IRLM

As use of the IRLM for different study designs and purposes continues to expand and evolve, we envision supporting researchers and other program implementers in applying the IRLM to their own contexts. Our team at Northwestern University hosts web resources on the IRLM that includes completed examples and tools to assist users in completing their model, including templates in various formats (Additional Files A1– A5 and others) a Quick Reference Guide (Additional File A6) and a series of worksheets that provide guidance on populating the IRLM (Additional File A7). These will be available at https://cepim.northwestern.edu/implementationresearchlogicmodel/.

## Discussion

The IRLM provides a compact visual depiction of an implementation project and is a useful tool for academic–practice collaboration and partnership development. Its usability is high for seasoned and novice implementation researchers alike, as evidenced by our survey results and preliminary work. Its use in the planning, executing, reporting, and synthesizing of implementation research could increase the rigor and transparency of complex studies that ultimately could improve reproducibility—a challenge in the field—by offering a common structure to increase consistency and a method for more clearly specifying links and pathways to test theories.

Among the drawbacks of the IRLM is that it might be viewed as a somewhat simplified format. This represents the challenges of balancing depth and detail with parsimony, ease of comprehension, and ease of use. The structure of the IRLM may inhibit creative thinking if applied too rigidly, which is among the reasons we provide numerous examples of different ways to tailor the model to the specific needs of different project designs and parameters. Relatedly, we encourage users to iterate on the design of the IRLM to increase its utility.

## Conclusions

The promise of implementation science lies in the ability to conduct rigorous and reproducible research, to clearly understand the findings, and to synthesize findings from which generalizable conclusions can be drawn and actionable recommendations for practice change emerge. As scientists and implementers have worked to better define the core methods of the field, the need for theory-driven, testable integration of the foundational elements involved in impactful implementation research has become more apparent. The IRLM is a tool that can aid the field in addressing this need and moving toward the ultimate promise of implementation research: to improve the provision and quality of healthcare services for all people.

## Data Availability

Not applicable.

## List of Abbreviations

CFIR: Consolidated Framework for Implementation Research
EBP: Evidence-based practice
ERIC: Expert Recommendations for Implementing Change
IRLM: Implementation Research Logic Model
ISC^3^i: Implementation Science Coordination, Consultation, and Collaboration

## Declarations

### Ethics approval and consent to participate

Not applicable. This study did not involve human subjects.

### Availability of data and material

Not applicable.

### Competing interests

None declared.

### Funding

This study was supported by grant P30 DA027828 from the National Institute on Drug Abuse, awarded to C. Hendricks Brown; grant U18 DP006255 to Justin Smith and Cady Berkel; grant R56 HL148192 to Justin Smith; grant UL1 TR001422 from the National Center for Advancing Translational Sciences to Donald Lloyd-Jones; grant R01 MH118213 to Brian Mustanski; grant P30 AI117943 from the National Institute of Allergy and Infectious Diseases to Richard D’Aquila; grant UM1 CA233035 from the National Cancer Institute to David Cella; a grant from the Woman’s Board of Northwestern Memorial Hospital to John Csernansky; grant F32 HS025077 from the Agency for Healthcare Research and Quality; grant NIFTI 2016-20178 from the Foundation for Physical Therapy; the Shirley Ryan AbilityLab; and by the Implementation Research Institute (IRI) at the George Warren Brown School of Social Work, Washington University in St. Louis through grant R25 MH080916 from the National Institute of Mental Health and the Department of Veterans Affairs, Health Services Research & Development Service, Quality Enhancement Research Initiative (QUERI) to Enola Proctor. The opinions expressed herein are the views of the authors and do not necessarily reflect the official policy or position of the National Institutes of Health, the Centers for Disease Control and Prevention, the Agency for Healthcare Research and Quality the Department of Veterans Affairs..

### Authors’ contributions

JDS conceived of the Implementation Research Logic Model. JDS, MR, and DL collaborated in developing the Implementation Research Logic Model as presented and in the writing of the manuscript. All authors approved of the final version.

## Acknowledgements

The authors wish to thank colleagues who provided input at different stages of developing this manuscript and the Implementation Research Logic Model: Hendricks Brown, Brian Mustanski, Kathryn Macapagal, Nanette Benbow, Lisa Hirschhorn, Richard Lieber, Piper Hansen, Leslie O’Donnell, Allen Heinemann, Enola Proctor, Courtney Wolk-Benjamin, Sandra Naoom, Emily Fu, Jeffrey Rado, Lisa Rosenthal, Patrick Sullivan, Aaron Siegler, Cady Berkel, Carrie Dooyema, Lauren Fiechtner, Jeanne Lindros, Vinny Biggs, Gerri Cannon-Smith, Jeremiah Salmon, Sujata Ghosh, Alison Baker.

## Legend

**Additional file 1**

IRLM Fillable PDF form

**Additional file 2**

IRLM for Comparative Implementation

**Additional file 3**

IRLM for Implementation of an Intervention Across or Linking Two Contexts

**Additional file 4**

IRLM for an Implementation Optimization Study

**Additional file 5**

IRLM with Clinical or Preventive Intervention Specified

**Additional file 6**

IRLM Quick Reference Guide

**Additional file 7**

IRLM Worksheets

---

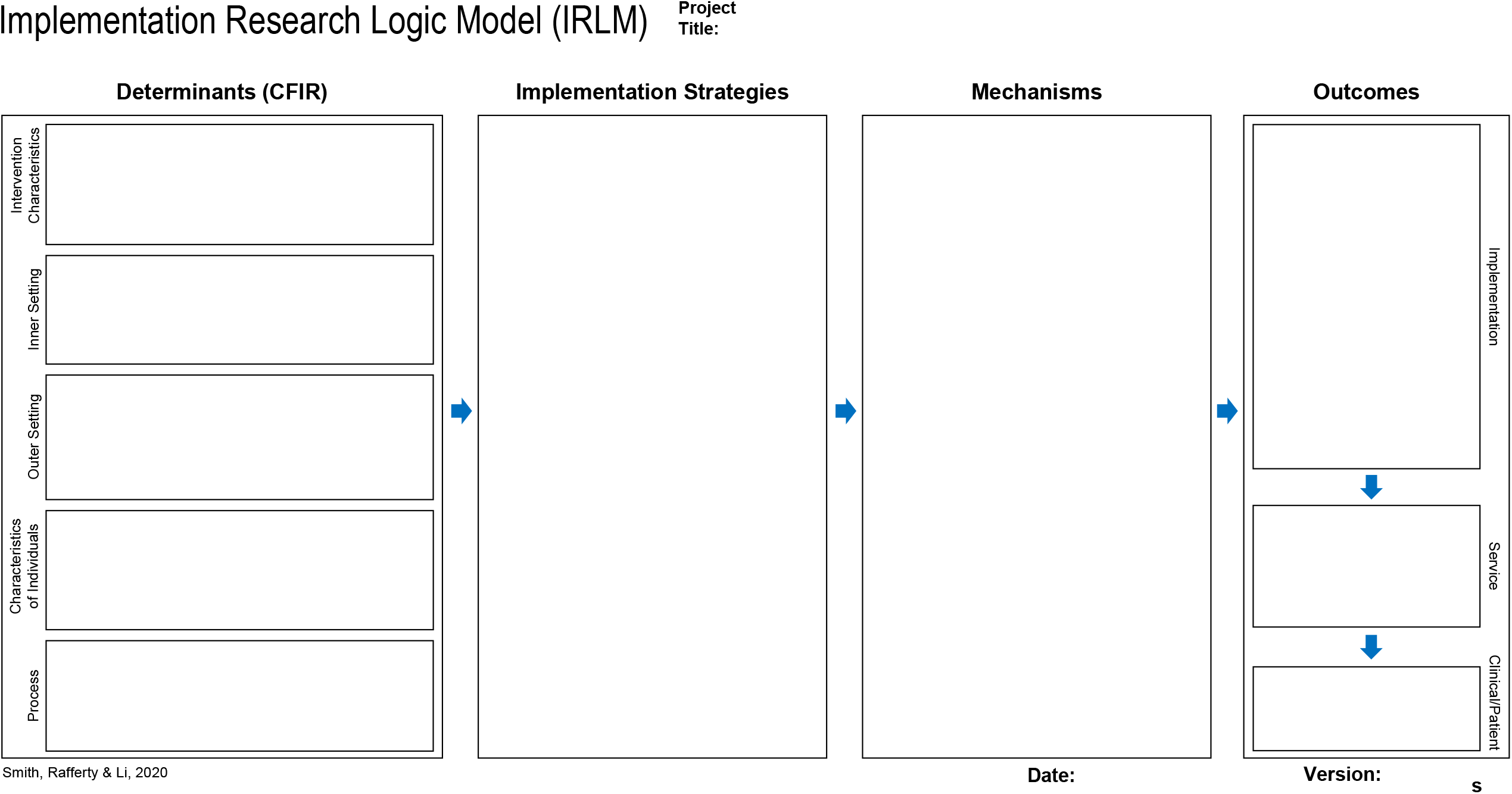

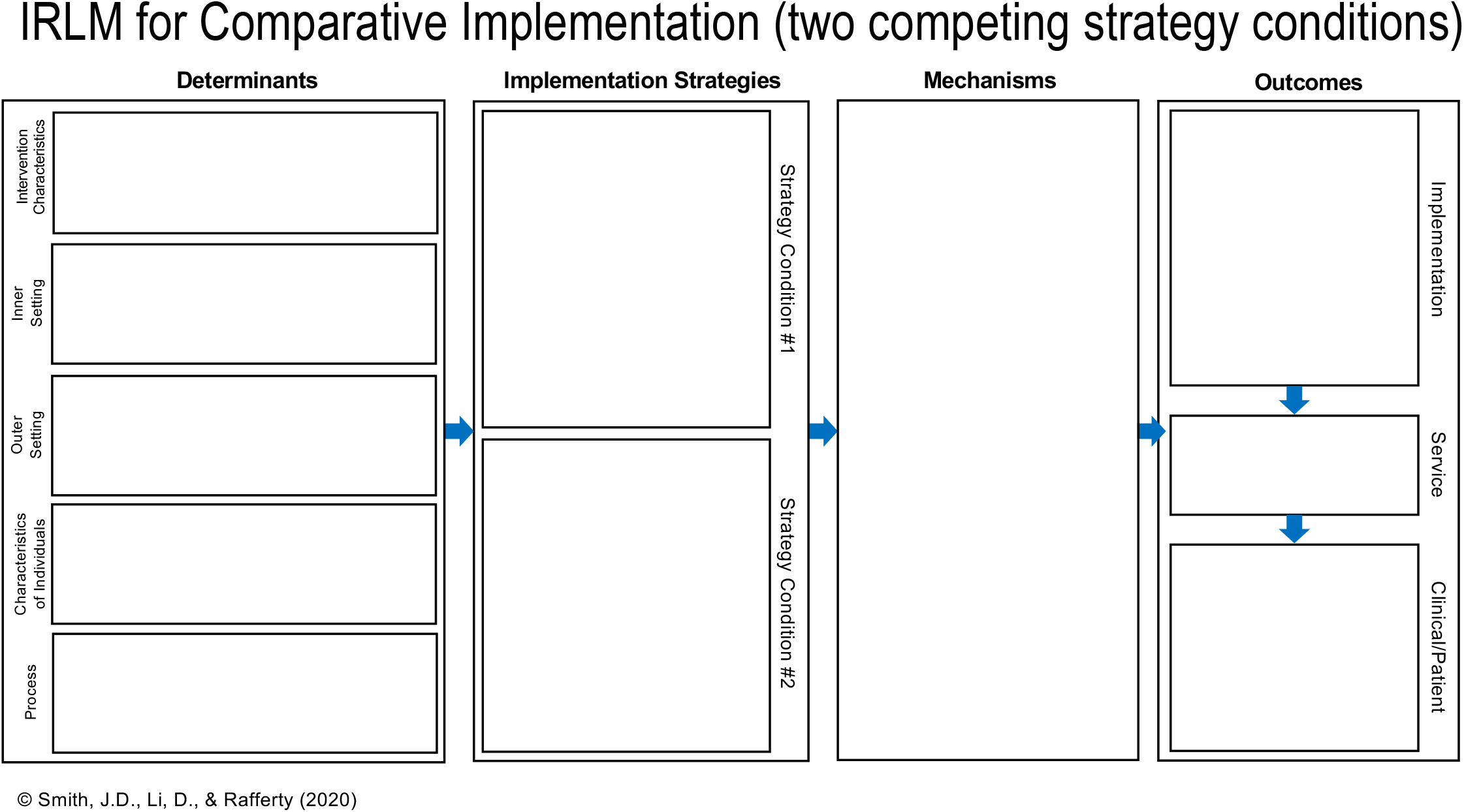

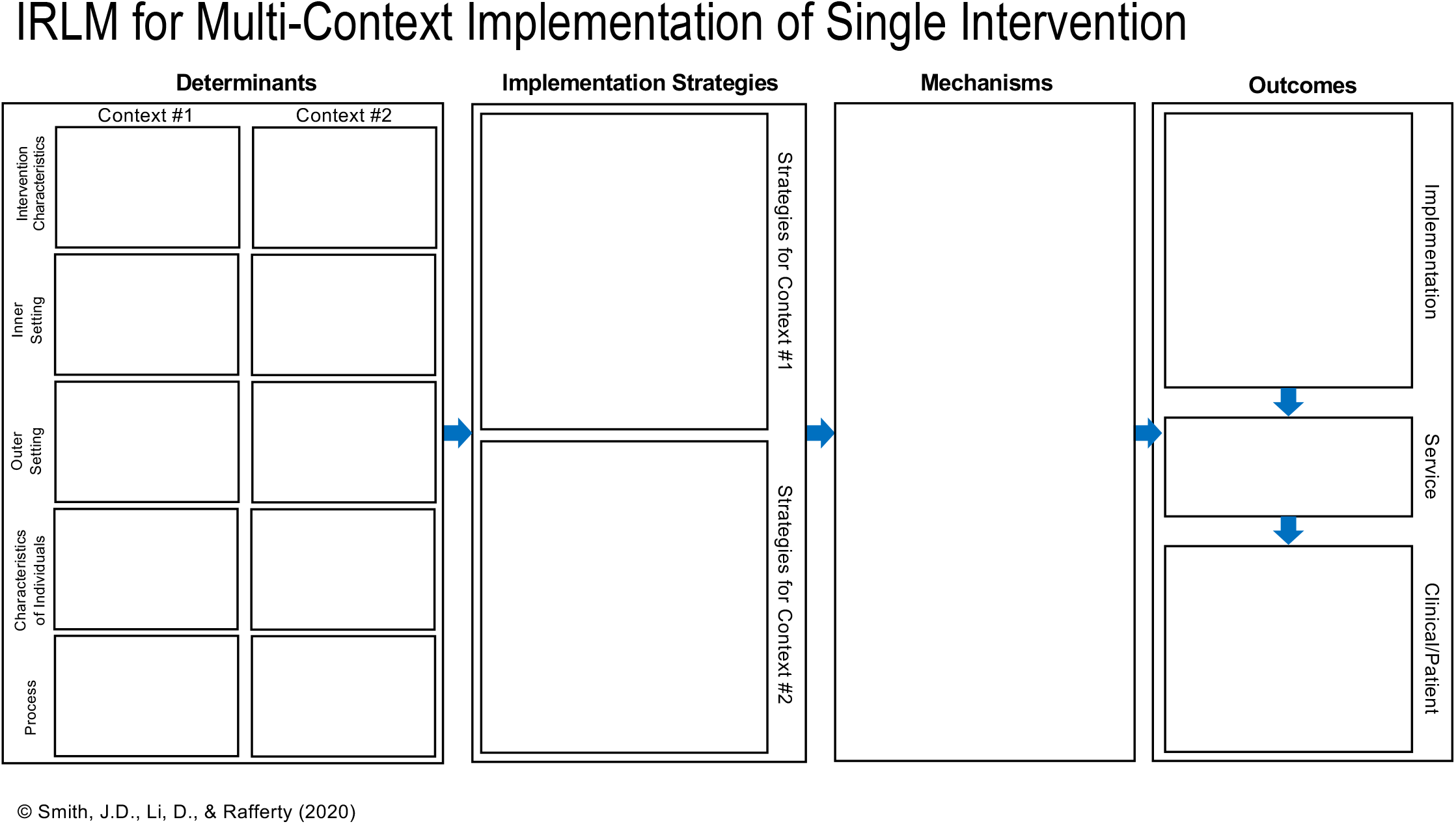

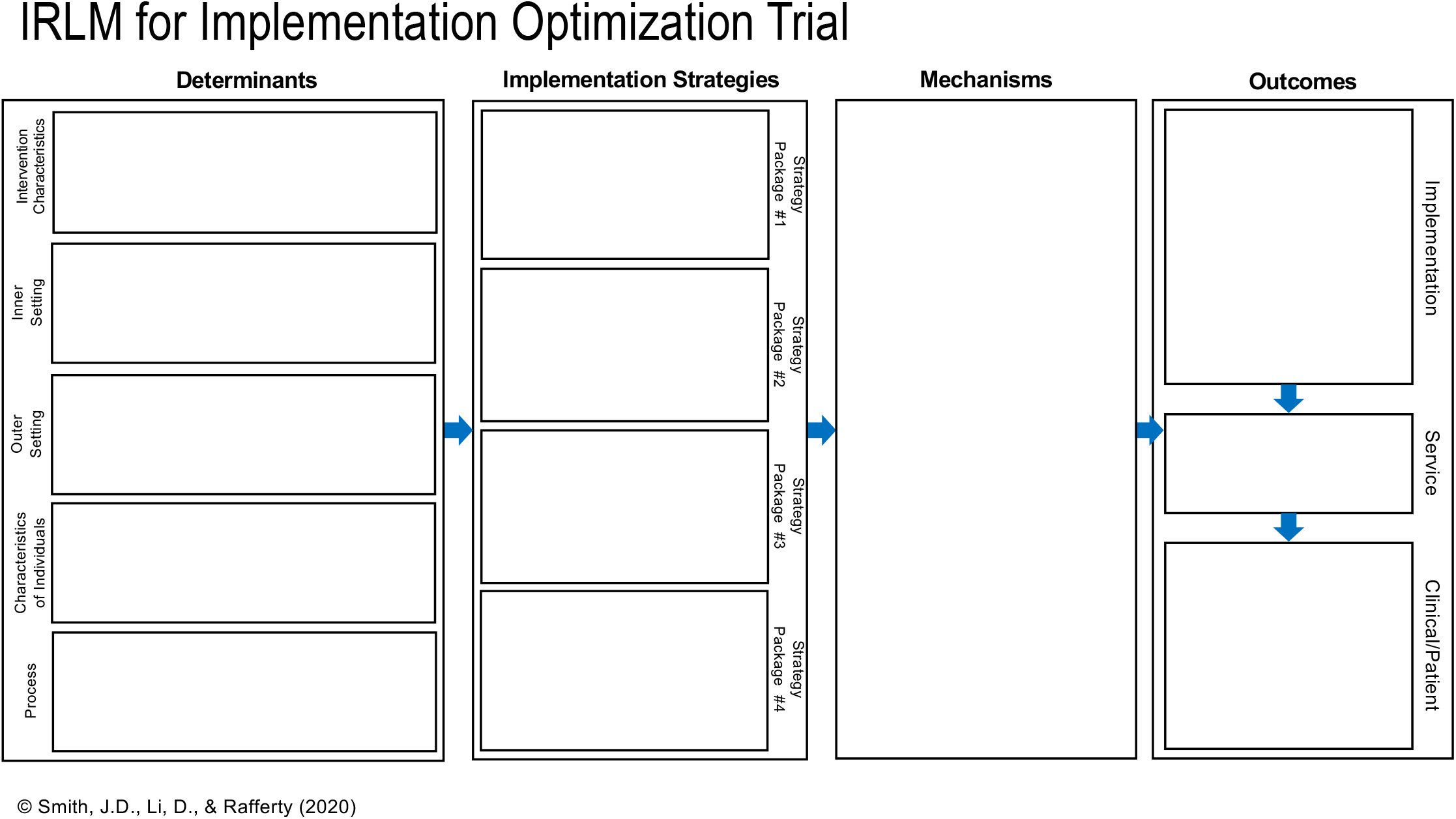

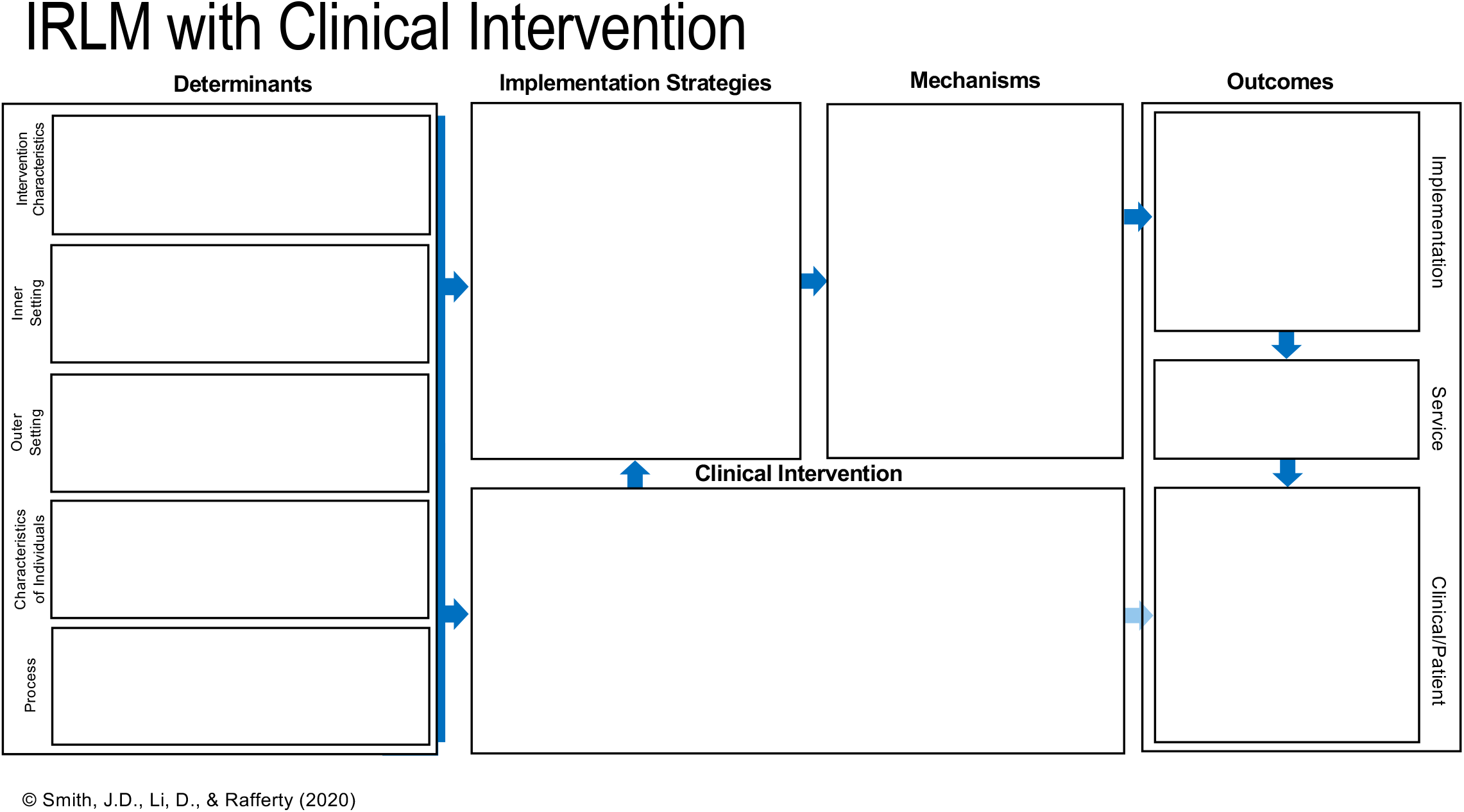

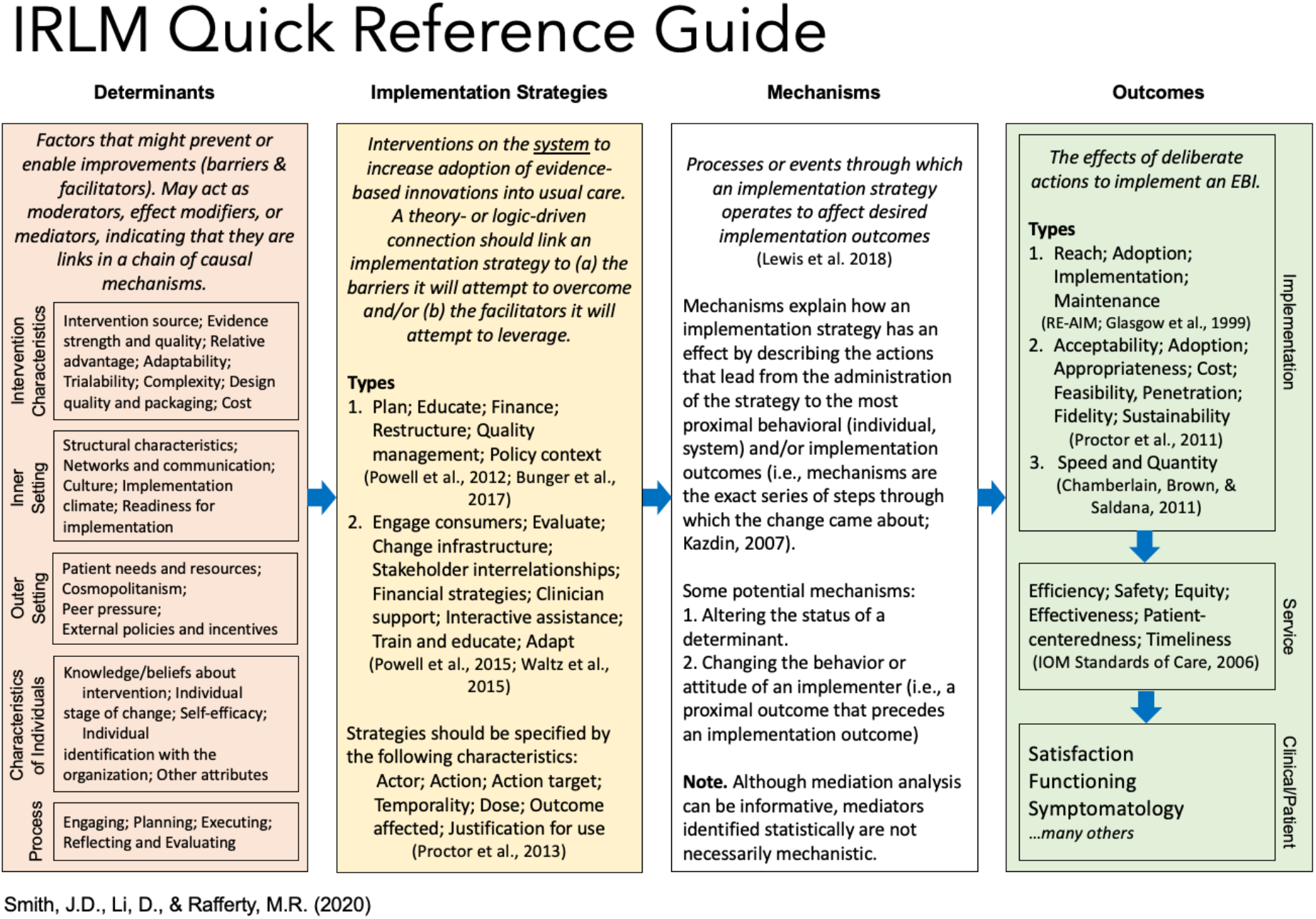

Determinants of implementation are constructs that have been associated with effective implementation. Often, researchers think of determinants as implementation barriers and facilitators, but they can also be mediators, moderators, predictors, and/or outcomes. One of the most comprehensive lists of determinants comes from the Consolidated Framework for Implementation Research (CFIR; Damschroder et al., 2009).

1. From the list of CFIR constructs below, place a checkmark (√) next to ones that may be germane to your project. It is important to capture all factors that may affect the implementation of your intervention.
2. Circle any determinants that your project may aim to change/alter.
3. For each determinant, operationalize it for your project and add it to your **IRLM**.

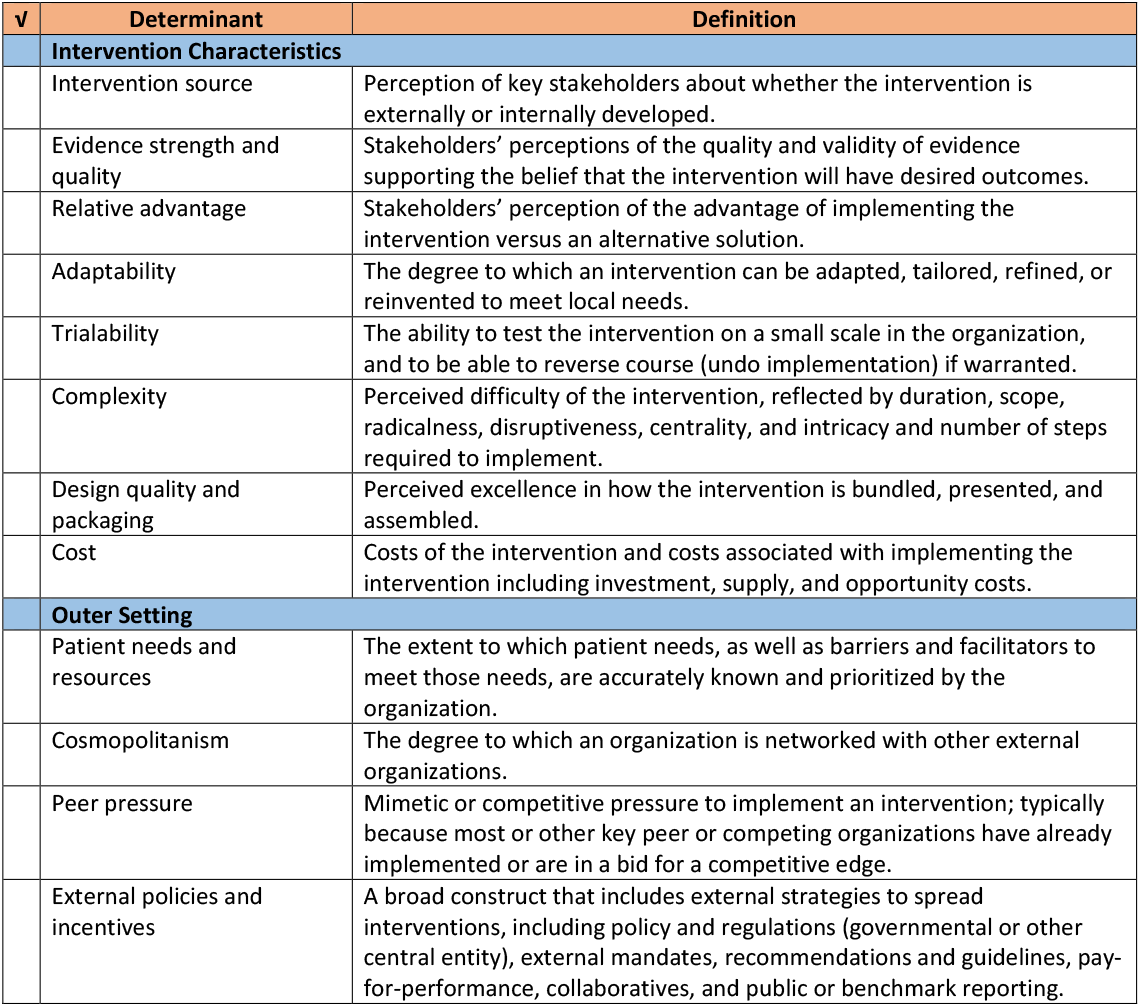
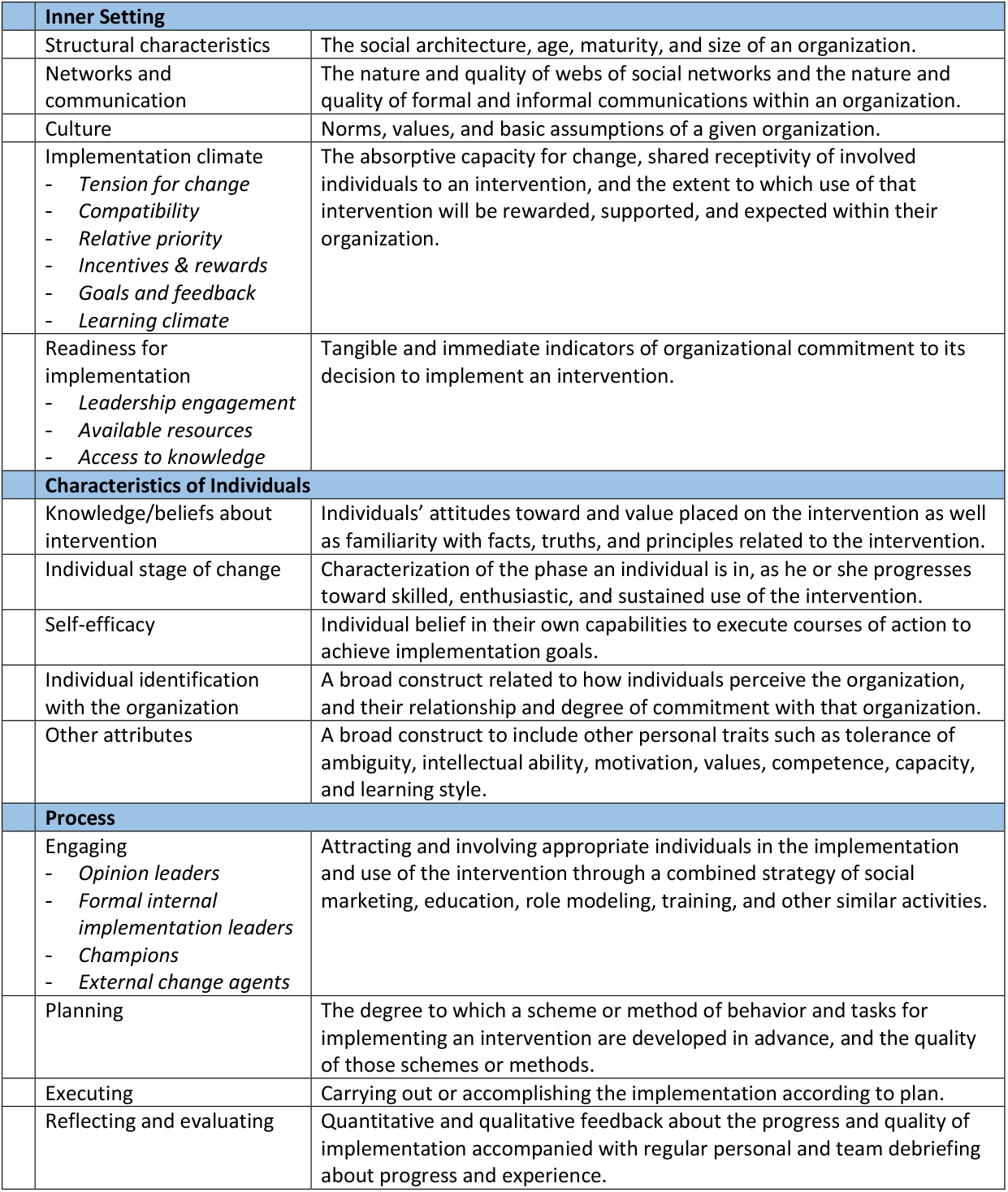

| ✓ | Determinant | Definition |
| --- | --- | --- |
| **Intervention Characteristics** |  |  |
|  | Intervention source | Perception of key stakeholders about whether the intervention is externally or internally developed. |
|  | Evidence strength and quality | Stakeholders' perceptions of the quality and validity of evidence supporting the belief that the intervention will have desired outcomes. |
|  | Relative advantage | Stakeholders' perception of the advantage of implementing the intervention versus an alternative solution. |
|  | Adaptability | The degree to which an intervention can be adapted, tailored, refined, or reinvented to meet local needs. |
|  | Trialability | The ability to test the intervention on a small scale in the organization, and to be able to reverse course (undo implementation) if warranted. |
|  | Complexity | Perceived difficulty of the intervention, reflected by duration, scope, radicalness, disruptiveness, centrality, and intricacy and number of steps required to implement. |
|  | Design quality and packaging | Perceived excellence in how the intervention is bundled, presented, and assembled. |
|  | Cost | Costs of the intervention and costs associated with implementing the intervention including investment, supply, and opportunity costs. |
| **Outer Setting** |  |  |
|  | Patient needs and resources | The extent to which patient needs, as well as barriers and facilitators to meet those needs, are accurately known and prioritized by the organization. |
|  | Cosmopolitanism | The degree to which an organization is networked with other external organizations. |
|  | Peer pressure | Mimetic or competitive pressure to implement an intervention; typically because most or other key peer or competing organizations have already implemented or are in a bid for a competitive edge. |
|  | External policies and incentives | A broad construct that includes external strategies to spread interventions, including policy and regulations (governmental or other central entity), external mandates, recommendations and guidelines, pay-for-performance, collaboratives, and public or benchmark reporting. |

| Inner Setting |  |
| --- | --- |
| Structural characteristics | The social architecture, age, maturity, and size of an organization. |
| Networks and communication | The nature and quality of webs of social networks and the nature and quality of formal and informal communications within an organization. |
| Culture | Norms, values, and basic assumptions of a given organization. |
| Implementation climate<br>- *Tension for change*<br>- *Compatibility*<br>- *Relative priority*<br>- *Incentives & rewards*<br>- *Goals and feedback*<br>- *Learning climate* | The absorptive capacity for change, shared receptivity of involved individuals to an intervention, and the extent to which use of that intervention will be rewarded, supported, and expected within their organization. |
| Readiness for implementation<br>- *Leadership engagement*<br>- *Available resources*<br>- *Access to knowledge* | Tangible and immediate indicators of organizational commitment to its decision to implement an intervention. |
| Characteristics of individuals |  |
| Knowledge/beliefs about intervention | Individuals' attitudes toward and value placed on the intervention as well as familiarity with facts, truths, and principles related to the intervention. |
| Individual stage of change | Characterization of the phase an individual is in, as he or she progresses toward skilled, enthusiastic, and sustained use of the intervention. |
| Self-efficacy | Individual belief in their own capabilities to execute courses of action to achieve implementation goals. |
| Individual identification with the organization | A broad construct related to how individuals perceive the organization, and their relationship and degree of commitment with that organization. |
| Other attributes | A broad construct to include other personal traits such as tolerance of ambiguity, intellectual ability, motivation, values, competence, capacity, and learning style. |
| Process |  |
| Engaging<br>- *Opinion leaders*<br>- *Formal internal implementation leaders*<br>- *Champions*<br>- *External change agents* | Attracting and involving appropriate individuals in the implementation and use of the intervention through a combined strategy of social marketing, education, role modeling, training, and other similar activities. |
| Planning | The degree to which a scheme or method of behavior and tasks for implementing an intervention are developed in advance, and the quality of those schemes or methods. |
| Executing | Carrying out or accomplishing the implementation according to plan. |
| Reflecting and evaluating | Quantitative and qualitative feedback about the progress and quality of implementation accompanied with regular personal and team debriefing about progress and experience. |

Implementation outcomes are “the effects of deliberate and purposive actions to implement new treatments, practices, and services” (Proctor et al., 2011). They serve as (1) indicators of implementation success, (2) proximal indicators of implementation processes, and (3) *intermediate outcomes in relation to service and clinical/patient outcomes*:

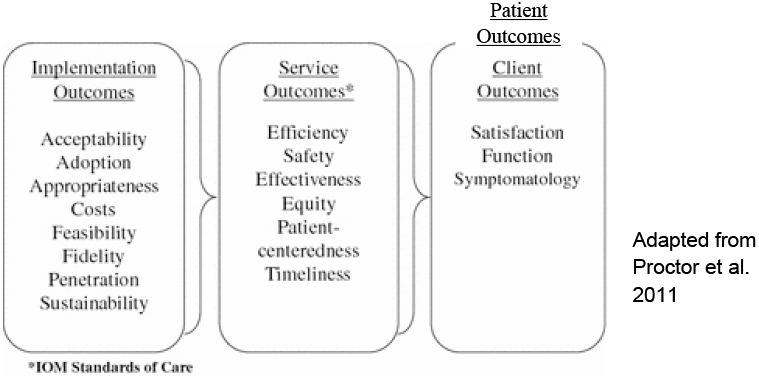

Unlike clinical/patient outcomes, implementation outcomes are often at the level of the system, setting, or service provider and typically not at the level of the patient/client. Some outcomes may be measured by researchers, whereas other may be measured through administrative records.

To identify implementation outcomes for your project, it is helpful to work backward from the most downstream/ distal/long-term to more upstream/proximal/short-term outcomes.

1. For the evidence-based intervention that is the focus of your project, what are the clinical/patient outcomes you are interested in? These may include clinical indicators, patient behaviors, patient-reported outcomes, etc. Add these to your **IRLM**.
2. From the list of service outcomes below, place a checkmark (√) next to ones that may be relevant to your project. Add these to your **IRLM**.

3. From the list of implementation outcomes below, place a checkmark (√) next to ones that may be germane to your project. For each outcome, operationalize it for your project and add it to your **IRLM**.

| ✓ | Service outcome | Definition |
| --- | --- | --- |
|  | Efficiency | Avoiding waste, including waste of equipment, supplies, ideas, and energy. |
|  | Safety | Avoiding harm to patients from the care that is intended to help them. |
|  | Effectiveness | Providing services based on scientific knowledge to all who could benefit and refraining from providing services to those not likely to benefit (avoiding underuse and misuse, respectively). |
|  | Equity | Providing care that does not vary in quality because of personal characteristics such as gender, ethnicity, geographic location, and socioeconomic status. |
|  | Patient-centeredness | Providing care that is respectful of and responsive to individual patient preferences, needs, and values and ensuring that patient values guide all clinical decisions. |
|  | Timeliness | Reducing waits and sometimes harmful delays for both those who receive and those who give care. |

| ✓ | Implementation outcome | Definition |
| --- | --- | --- |
| **RE-AIM Framework (www.re-aim.org; Glasgow, Vogt, & Boles, 1999)** |  |  |
|  | Reach | The absolute number, proportion, and representativeness of individuals who are willing to participate in a given initiative, intervention, or program. |
|  | (Effectiveness) | (See service outcomes.) The impact of an intervention on important outcomes, including potential negative effects, quality of life, and economic outcomes. |
|  | Adoption | The absolute number, proportion, and representativeness of settings and intervention agents (people who deliver the program) who are willing to initiate a program. |
|  | Implementation | At the setting level, implementation refers to the intervention agents' fidelity to the various elements of an intervention's protocol, including consistency of delivery as intended and the time and cost of the intervention. At the individual level, implementation refers to clients' use of the intervention strategies. |
|  | Maintenance | The extent to which a program or policy becomes institutionalized or part of the routine organizational practices and policies. Within the RE-AIM framework, maintenance also applies at the individual level. At the individual level, maintenance has been defined as the long-term effects of a program on outcomes after 6 or more months after the most recent intervention contact. |
| **Proctor et al., 2011** |  |  |
|  | Acceptability | The perception among implementation stakeholders that a given treatment, service, practice, or innovation is agreeable, palatable, or satisfactory. |
|  | Adoption | The intention, initial decision, or action to try or employ an innovation or evidence-based practice. |
|  | Appropriateness | The perceived fit, relevance, or compatibility of the innovation or evidence based practice for a given practice setting, provider, or consumer; and/or perceived fit of the innovation to address a particular issue or problem. |
|  | Cost | The cost impact of an implementation effort. |
|  | Feasibility | The extent to which a new treatment, or an innovation, can be successfully used or carried out within a given agency or setting. |
|  | Fidelity | The degree to which an intervention was implemented as it was prescribed in the original protocol or as it was intended by the program developers. |
|  | Penetration/Uptake | The integration of a practice within a service setting and its subsystems. |
|  | Sustainability | The extent to which a newly implemented treatment is maintained or institutionalized within a service setting's ongoing, stable operation. |

In implementation research, the word “intervention” can refer to two things:

- An evidence-based intervention→ the program, policy, practice, pill, etc., that affects patient outcomes.
- An implementation intervention→ manipulations to the *system* that help implement the EBI. To avoid inevitable confusion, we typically refer to the latter as “strategies.”

When implementing an EBI, multiple discrete strategies are typically used. Several taxonomies/lists of strategies exist in the literature, including by Bunger et al. (2017) and by Powell et al. (2015).

1. From either taxonomy below, place a checkmark (√) next to strategy categories that you may be considering for your project.
  a. For help selecting strategies based on your determinants of implementation, you may use the CFIR- ERIC Matching Tool found at https://cfirguide.org/choosing-strategies/.
2. For each strategy category, identify discrete strategies and operationalize them for your project.
  a. A full list of Bunger et al. strategies can be found at https://link.springer.com/article/10.1186/s12961-017-0175-y.
  b. A full list of the Powell et al. (a.k.a. ERIC) strategies can be found at https://implementationscience.biomedcentral.com/articles/10.1186/s13012-015-0295-0/tables/1.
3. Add your discrete strategies to your logic model. See the completed logic model for examples. E.g., the PrEP example project used *education* (Bunger)/ *train and educate stakeholders* (ERIC) to train providers/staff on PrEP efficacy, eligibility, stigma, etc.

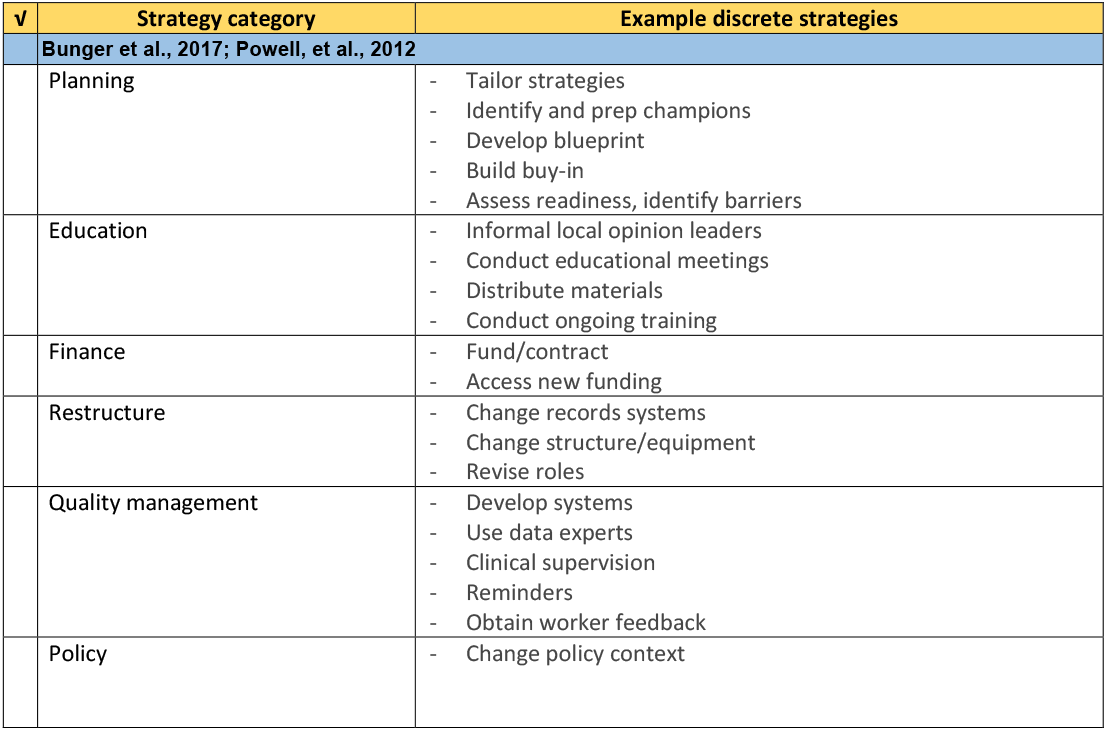
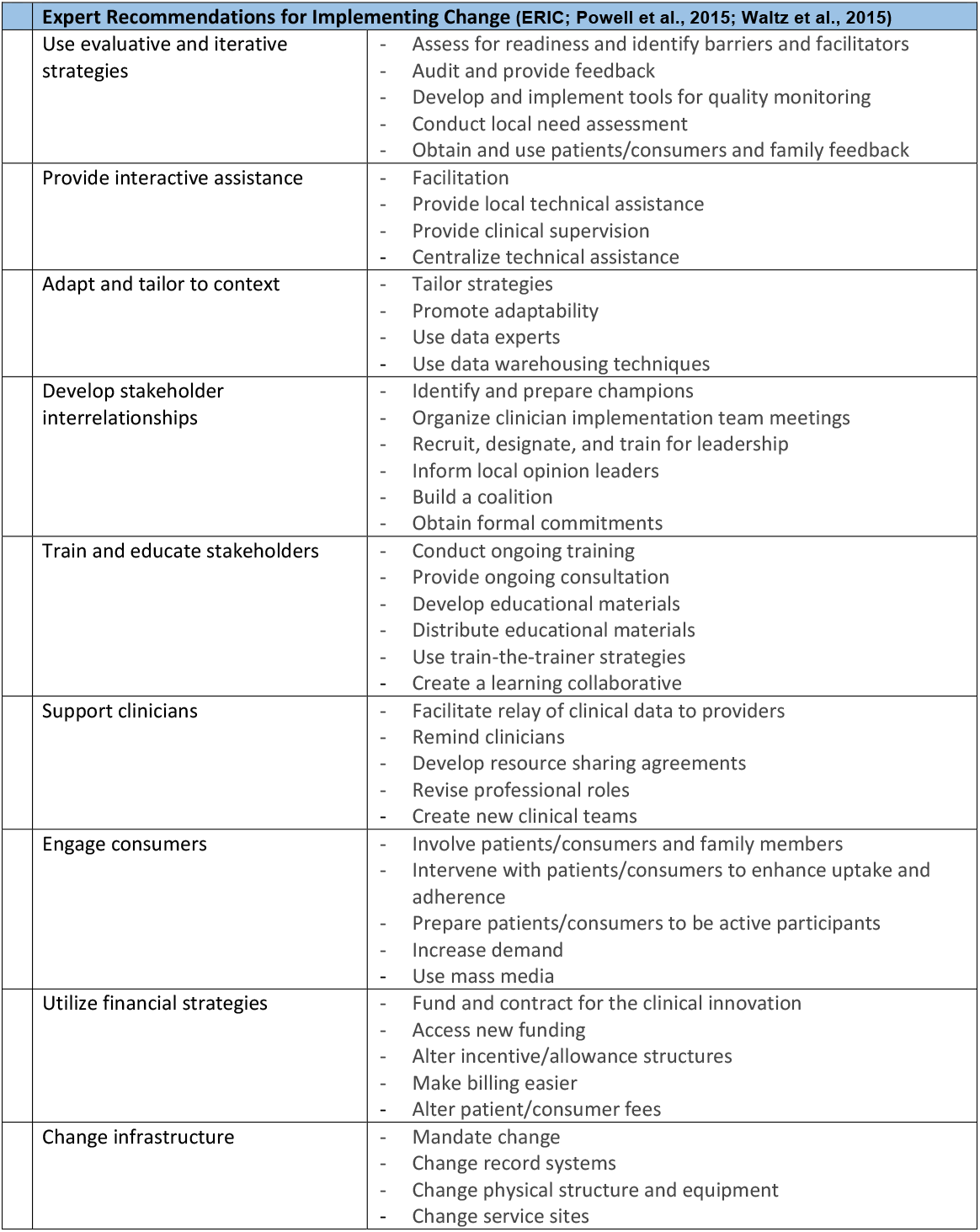

## Notes

### Competing Interest Statement

The authors have declared no competing interest.

## References

1. Nosek BA, Alter G, Banks GC, Borsboom D, Bowman SD, Breckler SJ, Buck S, Chambers CD, Chin G, Christensen G, et al: Promoting an open research culture. Science 2015, 348:1422–1425.

2. Slaughter SE, Hill JN, Snelgrove-Clarke E: What is the extent and quality of documentation and reporting of fidelity to implementation strategies: a scoping review. Implementation Science 2015, 10:1–12.

3. Brown CH, Curran G, Palinkas LA, Aarons GA, Wells KB, Jones L, Collins LM, Duan N, Mittman BS, Wallace A, et al: An overview of research and evaluation designs for dissemination and implementation. Annual Review of Public Health 2017, 38:null.

4. Hwang S, Birken SA, Melvin CL, Rohweder CL, Smith JD: Designs and methods for implementation research: Advancing the mission of the CTSA program. Journal of Clinical and Translational Science 2020:Available online.

5. Smith JD: An implementation research logic model: A step toward improving scientific rigor, transparency, reproducibility, and specification. Implementation Science 2018, 14:S39.

6. Tabak RG, Khoong EC, Chambers DA, Brownson RC: Bridging research and practice: Models for dissemination and implementation research. American Journal of Preventive Medicine 2012, 43:337–350.

7. Nilsen P: Making sense of implementation theories, models and frameworks. Implementation Science 2015, 10:53.

8. Damschroder LJ: Clarity out of chaos: Use of theory in implementation research. Psychiatry Research 2019.

9. Proctor EK, Powell BJ, McMillen JC: Implementation strategies: recommendations for specifying and reporting. Implement Sci 2013, 8.

10. Kessler RS, Purcell EP, Glasgow RE, Klesges LM, Benkeser RM, Peek CJ: What Does It Mean to “Employ” the RE-AIM Model? Evaluation & the Health Professions 2013, 36:44–66.

11. Pinnock H, Barwick M, Carpenter CR, Eldridge S, Grandes G, Griffiths CJ, Rycroft-Malone J, Meissner P, Murray E, Patel A, et al: Standards for Reporting Implementation Studies (StaRI): explanation and elaboration document. BMJ Open 2017, 7:e013318.

12. Lewis CC, Klasnja P, Powell BJ, Lyon AR, Tuzzio L, Jones S, Walsh-Bailey C, Weiner B: From Classification to Causality: Advancing Understanding of Mechanisms of Change in Implementation Science. Frontiers in Public Health 2018, 6.

13. Glanz K, Bishop DB: The Role of Behavioral Science Theory in Development and Implementation of Public Health Interventions. Annual Review of Public Health 2010, 31:399–418.

14. WK Kellogg Foundation: Logic model development guide. Battle Creek, Michigan: WK Kellogg Foundation; 2004.

15. CFAR/ARC Ending the HIV Epidemic Supplement Awards [https://www.niaid.nih.gov/research/cfar-arc-ending-hiv-epidemic-supplement-awards]

16. Funnell SC, Rogers PJ: Purposeful program theory: Effective use of theories of change and logic models. San Francisco, CA: John Wiley & Sons; 2011.

17. Petersen D, Taylor EF, Peikes D: The Logic Model: The Foundation to Implement, Study, and Refine Patient-Centered Medical Home Models (Issue Brief). Mathematica Policy Research Reports. Mathematica Policy Research; 2013.

18. Fernandez ME, ten Hoor GA, van Lieshout S, Rodriguez SA, Beidas RS, Parcel G, Ruiter RAC, Markham CM, Kok G: Implementation Mapping: Using Intervention Mapping to Develop Implementation Strategies. Frontiers in Public Health 2019, 7.

19. Proctor EK, Landsverk J, Aarons G, Chambers D, Glisson C, Mittman B: Implementation research in mental health services: An emerging science with conceptual, methodological, and training challenges. Adm Policy Ment Health 2009, 36.

20. Proctor EK, Silmere H, Raghavan R, Hovmand P, Aarons G, Bunger A, Griffey R, Hensley M: Outcomes for implementation research: conceptual distinctions, measurement challenges, and research agenda. Adm Policy Ment Health Ment Health Serv Res 2011, 38.

21. Smith JD, Rafferty MR, Heinemann AW, Meachum MK, Villamar JA, Lieber RL, Brown CH: Evaluation of the factor structure of implementation research measures adapted for a novel context and multiple professional roles. BMC Health Serv Res 2020.

22. Smith JD, Fu E, Carroll AJ, Rado J, Rosenthal LJ, Atlas JA, Burnett-Zeigler I, Jordan N, Brown CH, Csernansky J: Collaborative Care for DepressionManagement in Primary Care: A Randomized Rollout Trial Using a Type 2 Hybrid Effectiveness-Implementation Design submitted for publication.

23. Smith JD, Berkel C, Jordan N, Atkins DC, Narayanan SS, Gallo C, Grimm KJ, Dishion TJ, Mauricio AM, Rudo-Stern J, et al: An individually tailored family-centered intervention for pediatric obesity in primary care: Study protocol of a randomized type II hybrid implementation-effectiveness trial (Raising Healthy Children study). Implementation Science 2018, 13:1–15.

24. Fauci AS, Redfield RR, Sigounas G, Weahkee MD, Giroir BP: Ending the HIV Epidemic: A Plan for the United States: Editorial. JAMA 2019, 321:844–845.

25. Rabin BA, Brownson RC: Terminology for dissemination and implementation research. In Dissemination and Implementation Research in Health 2nd ed. Edited by Brownson RC, Colditz G, Proctor EK. New York, NY: Oxford University Press; 2017: 19–46

26. Grimshaw JM, Eccles MP, Lavis JN, Hill SJ, Squires JE: Knowledge translation of research findings. Implementation Science 2012, 7:50.

27. Krause J, Van Lieshout J, Klomp R, Huntink E, Aakhus E, Flottorp S, Jaeger C, Steinhaeuser J, Godycki-Cwirko M, Kowalczyk A, et al: Identifying determinants of care for tailoring implementation in chronic diseases: an evaluation of different methods. Implementation Science 2014, 9:102.

28. Waltz TJ, Powell BJ, Fernández ME, Abadie B, Damschroder LJ: Choosing implementation strategies to address contextual barriers: diversity in recommendations and future directions. Implementation Science 2019, 14:42.

29. Damschroder LJ, Aron DC, Keith RE, Kirsh SR, Alexander JA, Lowery JC: Fostering implementation of health services research findings into practice: a consolidated framework for advancing implementation science. Implement Sci 2009, 4.

30. Atkins L, Francis J, Islam R, O’Connor D, Patey A, Ivers N, Foy R, Duncan EM, Colquhoun H, Grimshaw JM, et al : A guide to using the Theoretical Domains Framework of behaviour change to investigate implementation problems. Implementation Science 2017, 12:77.

31. Powell BJ, Waltz TJ, Chinman MJ, Damschroder LJ, Smith JL, Matthieu MM, Proctor EK, Kirchner JE: A refined compilation of implementation strategies: results from the Expert Recommendations for Implementing Change (ERIC) project. Implement Sci 2015, 10.

32. Powell BJ, Fernandez ME, Williams NJ, Aarons GA, Beidas RS, Lewis CC, McHugh SM, Weiner BJ: Enhancing the Impact of Implementation Strategies in Healthcare: A Research Agenda. Frontiers in Public Health 2019, 7.

33. PAR-19-274: Dissemination and Implementation Research in Health (R01 Clinical Trial Optional) [https://grants.nih.gov/grants/guide/pa-files/PAR-19-274.html]

34. Edmondson D, Falzon L, Sundquist KJ, Julian J, Meli L, Sumner JA, Kronish IM: A systematic review of the inclusion of mechanisms of action in NIH-funded intervention trials to improve medication adherence. Behaviour Research and Therapy 2018, 101:12–19.

35. Gaglio B, Shoup JA, Glasgow RE: The RE-AIM Framework: A systematic review of use over time. American Journal of Public Health 2013, 103:e38–e46.

36. Glasgow RE, Harden SM, Gaglio B, Rabin B, Smith ML, Porter GC, Ory MG, Estabrooks PA: RE-AIM Planning and Evaluation Framework: Adapting to New Science and Practice With a 20-Year Review. Frontiers in Public Health 2019, 7.

37. Glasgow RE, Vogt TM, Boles SM: Evaluating the public health impact of health promotion interventions: The RE-AIM framework. American Journal of Public Health 1999, 89:1322–1327.

38. Damschroder LJ, Reardon CM, Sperber N, Robinson CH, Fickel JJ, Oddone EZ: Implementation evaluation of the Telephone Lifestyle Coaching (TLC) program: organizational factors associated with successful implementation. Translational Behavioral Medicine 2016, 7:233–241.

39. Bunger AC, Powell BJ, Robertson HA, MacDowell H, Birken SA, Shea C: Tracking implementation strategies: a description of a practical approach and early findings. Health Research Policy and Systems 2017, 15:15.

40. Boyd MR, Powell BJ, Endicott D, Lewis CC: A Method for Tracking Implementation Strategies: An Exemplar Implementing Measurement-Based Care in Community Behavioral Health Clinics. Behavior Therapy 2018, 49:525–537.

41. Brown CH, Kellam S, Kaupert S, Muthén B, Wang W, Muthén L, Chamberlain P, PoVey C, Cady R, Valente T, et al: Partnerships for the design, conduct, and analysis of effectiveness, and implementation research: Experiences of thePrevention Science and Methodology Group. Administration and Policy in Mental Health and Mental Health Services Research 2012, 39:301–316.

42. McNulty M, Smith JD, Villamar J, Burnett-Zeigler I, Vermeer W, Benbow N, Gallo C, Wilensky U, Hjorth A, Mustanski B, et al: Implementation Research Methodologies for Achieving Scientific Equity and Health Equity. In Ethnicity & disease, vol. 29. pp. 83–92; 2019:83-92.

43. Collins LM, Murphy SA, Strecher V: The multiphase optimization strategy (MOST) and the sequential multiple assignment randomized trial (SMART): new methods for more potent eHealth interventions. Am J Prev Med 2007, 32:S112–118.

44. Brown CH, Ten Have TR, Jo B, Dagne G, Wyman PA, Muthén B, Gibbons RD: Adaptive designs for randomized trials in public health. Annual Review of Public Health 2009, 30:1–25.

45. Smith JD: The Roll-Out Implementation Optimization Design: Integrating Aims of Quality Improvement and Implementation Sciences. Submitted for publication 2020.

46. Dziak JJ, Nahum-Shani I, Collins LM: Multilevel factorial experiments for developing behavioral interventions: power, sample size, and resource considerations. Psychological methods 2012, 17:153–175.

47. MacKinnon DP, Lockwood CM, Hoffman JM, West SG, Sheets V: A comparison of methods to test mediation and other intervening variable effects. Psychological Methods 2002, 7:83–104.

48. Graham ID, Tetroe J: Planned action theories. In Knowledge translation in health care: Moving from evidence to practice. Edited by Straus S, Tetroe J, Graham ID. Hoboken, NJ: Wiley-Blackwell; 2009

49. Moullin JC, Dickson KS, Stadnick NA, Rabin B, Aarons GA: Systematic review of the Exploration, Preparation, Implementation, Sustainment (EPIS) framework. Implementation Science 2019, 14:1.

50. Rycroft-Malone J: The PARIHS Framework—A framework for guiding the implementation of evidence-based practice. Journal of nursing care quality 2004, 19:297–304.

